# Vulnerability to rumors during the COVID-19 pandemic: Results of a national survey

**DOI:** 10.1101/2020.10.01.20205187

**Authors:** Victoria Jane En Long, Wei Shien Koh, Young Ern Saw, Jean CJ Liu

## Abstract

Amidst the COVID-19 pandemic, many rumors have emerged. Given prior research linking rumor exposure to mental well-being, we conducted a nation-wide survey to document the base rate of rumor exposure and factors associated with rumor vulnerability. Between March to July 2020, 1237 participants were surveyed on 5 widely-disseminated COVID-19 rumors (that drinking water frequently could be preventive, that eating garlic could be preventive, that the outbreak arose because of bat soup consumption, that the virus was created in an American lab, and that the virus was created in a Chinese lab). For each rumor, participants reported whether they had heard, shared or believed each rumor. Although most participants had been exposed to COVID-19 rumors, few shared or believed these. Sharing behaviors sometimes occurred in the absence of belief; however, education emerged as a protective factor for both sharing and belief. Together, our results suggest that campaigns targeting skills associated with higher education (e.g. epistemology) may prove more effective than counter-rumor messages.

**Highlights:**

- Prior studies linked exposure to COVID-19 rumors with poor mental health.
- In a community sample, most participants reported having heard rumors.
- Few participants shared or believed rumors.
- Sharing sometimes occurred in the absence of belief.
- More educated individuals believed and shared fewer rumors.

## 1. Introduction

The global outbreak of coronavirus disease 2019 (COVID-19) has come with increased psychological burden. In several meta-analyses, depression and anxiety symptoms have been found to be elevated amongst healthcare workers and the general population (da Silva and Neto, 2021; Pappa et al., 2020; Salari et al., 2020). Others have reported a higher incidence of stress-related symptoms or of post-traumatic stress disorder (Cooke et al., 2020; Torales et al., 2020). These findings highlight the urgent need to understand factors predicting psychiatric outcomes, allowing vulnerable individuals to be identified and interventions to be developed.

In terms of predictors, exposure to COVID-19 rumors has emerged as a risk factor for poor mental health (Gao et al., 2020; Liu and Tong, 2020; Vardanjani et al., 2020; Xiong et al., 2020). This has occurred against the backdrop of an “infodemic” – a surge of COVID-19 misinformation created and shared primarily via social media (World Health Organization, 2020). In particular, the fast-changing nature of the pandemic means that accurate information has not always been accessible (Cuan-Baltazar et al., 2020; Hou et al., 2020), resulting in uncertainties that have given rise to a large number of rumors (Depoux et al., 2020; Larson, 2018; Vosoughi et al., 2018).

To date, several publicly-available datasets have been analyzed to document the spread of rumors. For example, during the early stage of the pandemic (December to April 2020), search engine keywords reflected popular myths (Rovetta and Bhagavathula, 2020; Singh et al., 2020), with a large number of searches pertaining to alternative medicines that had been speculated to prevent COVID-19 (e.g., garlic, Chinese medicinal herbs, or the malaria medication chloroquine; Hou et al., 2020). On the social media platform Twitter, conspiratorial theories were posted regarding disease origins – suggesting, for example, that the virus had been developed as a bioweapon or had resulted from the introduction of 5G mobile networks (Abd-Alrazaq et al., 2020; Ahmed et al., 2020).

In turn, the spread of COVID-19 rumors has led to deleterious consequences. In Iran for example, a myth that alcohol consumption could prevent or treat COVID-19 resulted in over 700 deaths related to methanol poisoning, with deaths attributed to methanol poisoning exceeding those attributed to COVID-19 in some provinces (Aghababaeian et al., 2020). Returning to mental health outcomes, the extent to which an individual has been exposed to, has shared, or believed in COVID-19 rumors has also been found to predict anxiety symptoms (Liu and Tong, 2020).

While demographic predictors of pandemic-related mental health are difficult to address (e.g., age, gender, pre-existing medical conditions; (Torales et al., 2020), a person’s exposure to rumors may constitute a modifiable risk factor (Abdoli, 2020; Ebrahim et al., 2020; Holmes et al., 2020). Correspondingly, efforts to develop interventions would benefit from an understanding of rumor vulnerability: (i) the base rates by which individuals are exposed to, believe in, or share rumors; and (ii) factors predicting these rumor-related experiences (Chua and Banerjee, 2018).

At present, little is known about individual vulnerability to COVID-19 rumors. While a handful of studies have surveyed individuals on their social media usage and enquired about rumor dissemination via these platforms (Banakar et al., 2020), we are not aware of any study that has identified persons most likely to encounter, believe in, or to share COVID-19 rumors. To address this gap in the literature, we thus conducted a nationwide survey examining rumor vulnerability during the COVID-19 pandemic.

## 2. Methods

### 2.1 Study design and population

Our study was conducted across five months in Singapore (7 March to 27 July 2020), a city-state in Asia that experienced a high number of COVID-19 cases in the early stage of the pandemic. During this time, we recruited 1237 participants who met the following eligibility criteria: (1) aged ≥21 years old, and (2) had lived in Singapore for ≥2 years. All participants were recruited via social media advertisements within community groups (e.g., groups for residential estates, universities, and workplaces), or through paid Facebook advertisements targeting Singapore-based users.

Upon study enrolment, participants provided informed consent and completed a 20-minute online survey via Qualtrics. As part of a larger study, participants reported their: (i) demographics, (ii) responses to the pandemic; (iii) sources of COVID-19 news; and (as we report in this paper) (iv) familiarity with rumors (Liu and Tong, 2020; Long and Liu, 2020; Saw et al., 2020). The study protocol was approved by the Yale-NUS College Ethics Review Committee (#2020-CERC-001) and was pre-registered on ClinicalTrials.gov (NCT04305574).

### 2.2 Outcome variables

As the primary outcome variables, we assessed participants’ familiarity with five rumors that had been widely spread during the COVID-19 pandemic: that (1) drinking water frequently will help prevent infection (COVID-19 prevention); (2) eating garlic can help prevent infection (COVID-19 prevention); (3) the outbreak arose from people eating bat soup (COVID-19 origins); (4) the virus was created in a US lab to affect China’s economy (COVID-19 origins); and (5) the virus was created in a Chinese lab as a bioweapon (COVID-19 origins). Rumors were selected for their widespread distribution both internationally and within the local context (Taylor, 2020; World Health Organisation, 2020).

For each rumor, participants indicated whether they: (1) had heard the claim before (yes/no); (2) thought the claim was true (yes/no); or (3) had shared the claim on social media (e.g., Facebook, WhatsApp) (yes/no). We assigned a score of 1 for “yes” responses, and summed across the rumors to create three scores: the total number of claims heard, the total number of claims believed, and the total number of claims shared. Finally, participants also indicated which of 13 possible sources they had encountered the rumors (e.g., Facebook, WhatsApp, online forums, television).

### 2.3 Predictor variables

As predictor variables, participants reported the following demographic details: age, gender, ethnicity, religion, country of birth, marital status, education, house type (a proxy of socio-economic status), and household size. Using the survey time-stamp, we also recorded two situational-related variables: the total number of local cases reported to date, and whether the country had been in a lock-down when participants completed the survey.

### 2.4 Statistical analysis

Using counts (%), we first summarized the baseline rates of rumor familiarity and rumor sources. As further exploratory analyses, we conducted Fisher’s exact test exploring the relationship between believing and sharing each rumor.

We then ran linear regression models to predict the following outcome measures: the total number of claims heard [Model 1], the total number of claims believed [Model 2], and the total number of claims shared [Model 3]. Each model involved the full set of predictor variables (described in 2.3), with the number of COVID-19 cases log-transformed for linearity.

For each model, we applied Bonferroni correction to control the type 1 family-wise error rate at 0.05 (Bonferroni-adjusted alpha level of 0.05/22 predictors = 0.002). All statistical analyses were conducted using R (Version 4.0).

## 3. Results

### 3.1 Response rate

Out of 1751 individuals who accessed the survey link, 1446 (82.6%) provided informed consent and participated in the survey. However, 209 (14.5%) participants did not complete the primary outcome measures (on COVID-19 rumors) and were excluded from statistical analyses.

The final sample of 1237 participants is comparable to the resident population with regards to: the proportion of participants born in Singapore, ethnicity, and household size (≤10% difference) and age. However, our sample had more participants who were female (63.9% vs. 51.1%), single (41.9% vs 18.8%), and university graduates (70.7% vs. 32.4%); and fewer participants who lived in 1-3 room public housing flats (8.2% vs. 23.7%) or who had Buddhist beliefs (14.6% vs 33.2%) (Table 1).

**Table 1.**
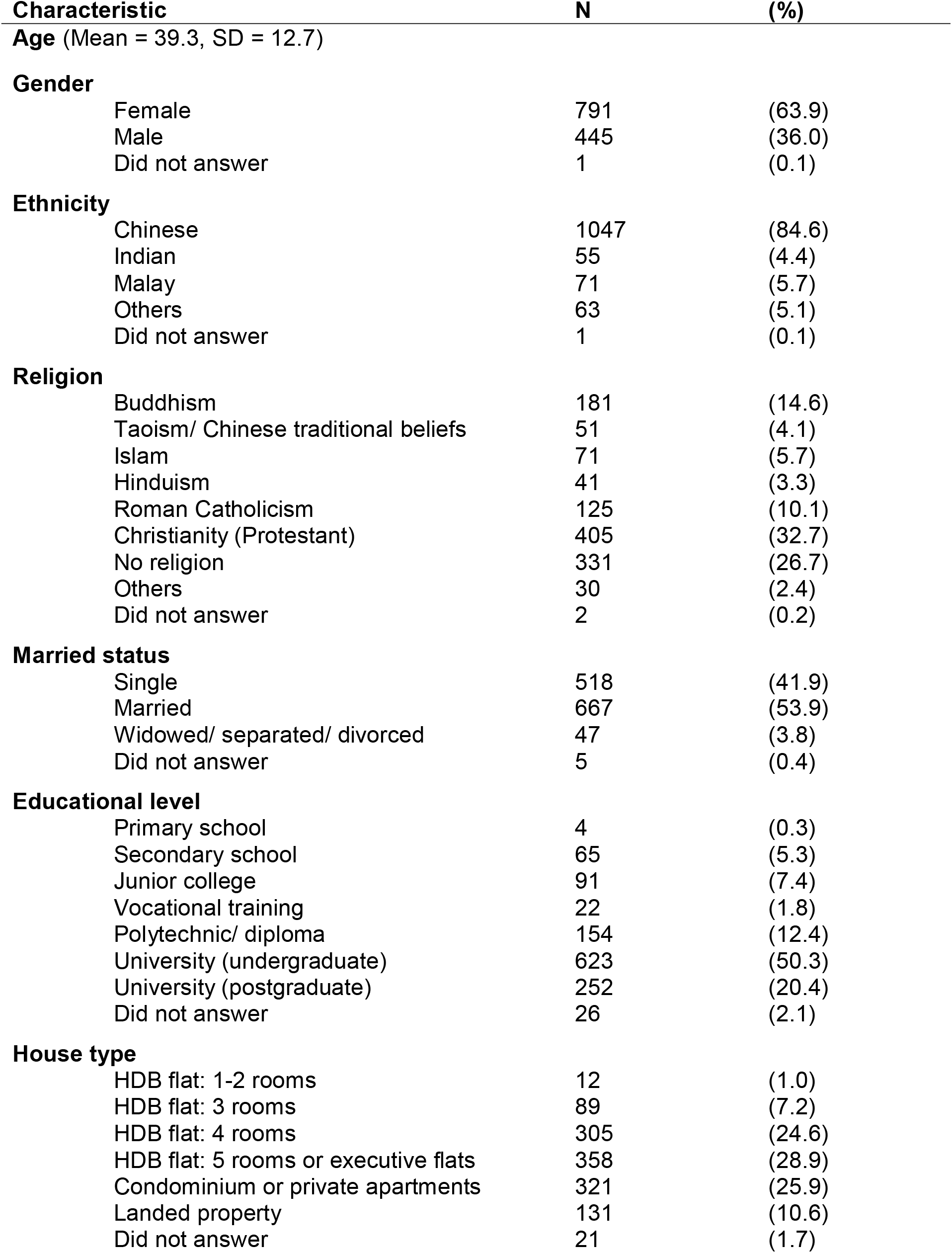
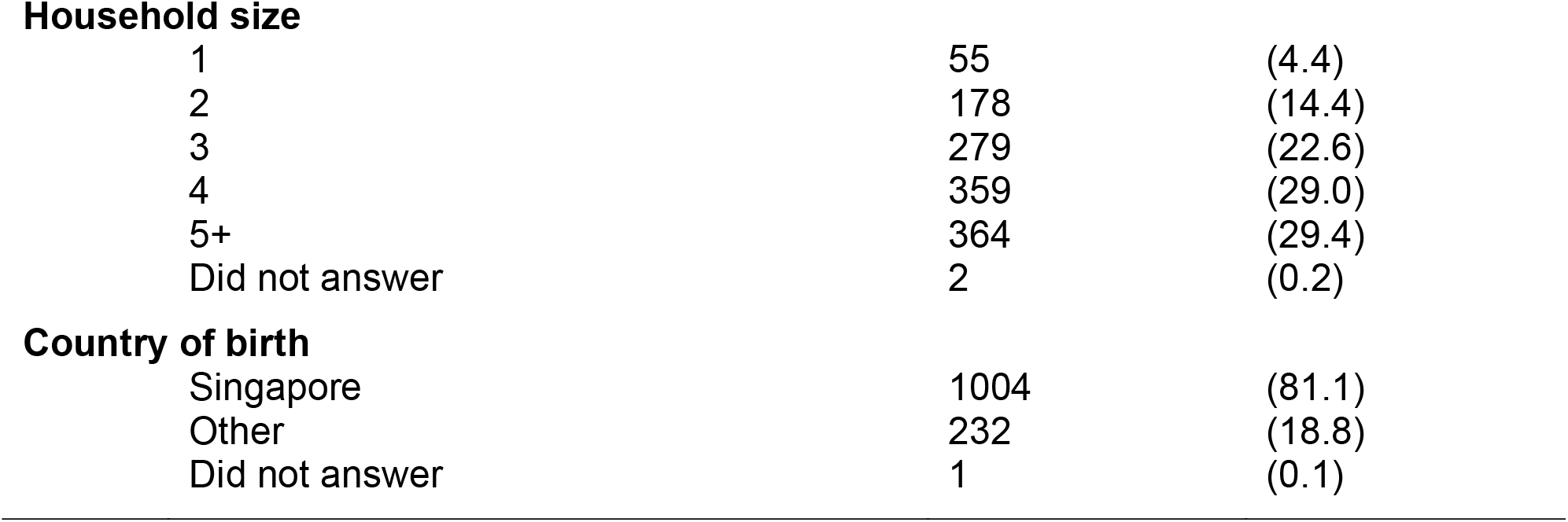
Baseline demographics of participants

| Characteristic | N | (%) |
| --- | --- | --- |
| **Age** (Mean = 39.3, SD = 12.7) |  |  |
| **Gender** |  |  |
| Female | 791 | (63.9) |
| Male | 445 | (36.0) |
| Did not answer | 1 | (0.1) |
| **Ethnicity** |  |  |
| Chinese | 1047 | (84.6) |
| Indian | 55 | (4.4) |
| Malay | 71 | (5.7) |
| Others | 63 | (5.1) |
| Did not answer | 1 | (0.1) |
| **Religion** |  |  |
| Buddhism | 181 | (14.6) |
| Taoism/ Chinese traditional beliefs | 51 | (4.1) |
| Islam | 71 | (5.7) |
| Hinduism | 41 | (3.3) |
| Roman Catholicism | 125 | (10.1) |
| Christianity (Protestant) | 405 | (32.7) |
| No religion | 331 | (26.7) |
| Others | 30 | (2.4) |
| Did not answer | 2 | (0.2) |
| **Married status** |  |  |
| Single | 518 | (41.9) |
| Married | 667 | (53.9) |
| Widowed/ separated/ divorced | 47 | (3.8) |
| Did not answer | 5 | (0.4) |
| **Educational level** |  |  |
| Primary school | 4 | (0.3) |
| Secondary school | 65 | (5.3) |
| Junior college | 91 | (7.4) |
| Vocational training | 22 | (1.8) |
| Polytechnic/ diploma | 154 | (12.4) |
| University (undergraduate) | 623 | (50.3) |
| University (postgraduate) | 252 | (20.4) |
| Did not answer | 26 | (2.1) |
| **House type** |  |  |
| HDB flat: 1-2 rooms | 12 | (1.0) |
| HDB flat: 3 rooms | 89 | (7.2) |
| HDB flat: 4 rooms | 305 | (24.6) |
| HDB flat: 5 rooms or executive flats | 358 | (28.9) |
| Condominium or private apartments | 321 | (25.9) |
| Landed property | 131 | (10.6) |
| Did not answer | 21 | (1.7) |
| **Household size** |  |  |
| 1 | 55 | (4.4) |
| 2 | 178 | (14.4) |
| 3 | 279 | (22.6) |
| 4 | 359 | (29.0) |
| 5+ | 364 | (29.4) |
| Did not answer | 2 | (0.2) |
| **Country of birth** |  |  |
| Singapore | 1004 | (81.1) |
| Other | 232 | (18.8) |
| Did not answer | 1 | (0.1) |

### 3.2 Base rates of familiarity with COVID-19 rumors

Out of 5 widely-disseminated rumors, the average participant had heard of 3.34 (*SD* = 1.33) rumors. The most commonly-heard rumor – reported by 8 in 10 participants (84.6%) – was that the outbreak had arisen from individuals eating bat soup. Despite high exposure to COVID-19 rumors, however, participants only believed an average of 0.27 claims (*SD* = 0.59) and shared 0.18 (*SD* = 0.63). The most commonly-believed rumor was that drinking water could prevent infection (11.4%), whereas the most commonly-shared rumor was again that the disease had arisen from bat soup consumption (7.1%) (Figure 1).

**Figure 1:**
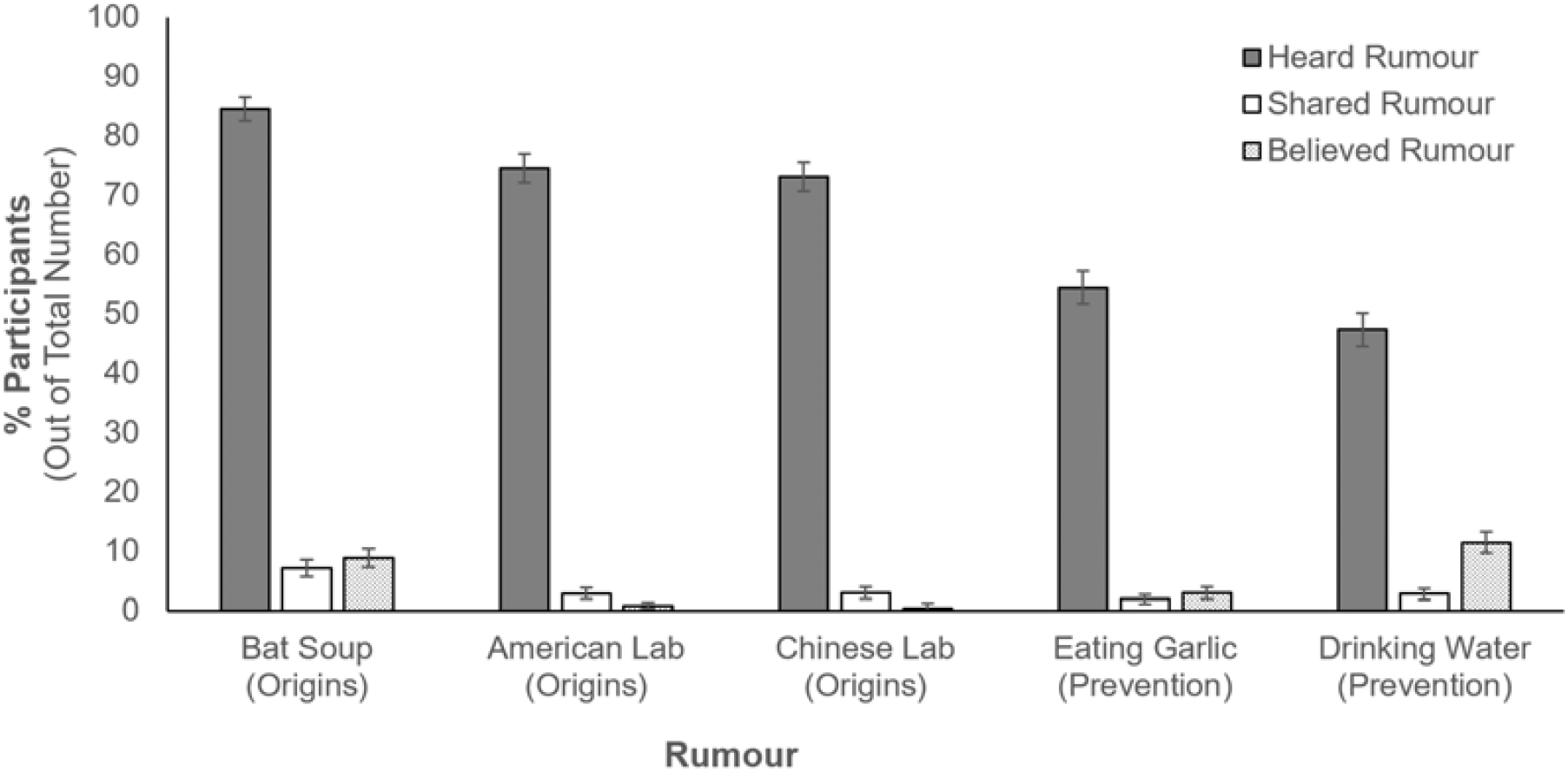
Proportion of participants hearing, sharing, and believe each COVID-19 rumor (that the virus originated from the consumption of bat soup, from an American lab, or from a Chinese lab; or that the virus can be cured by eating garlic or drinking water). Vertical lines represent the 95% confidence interval.)

In other words, as shown in Figure 2, most participants who had heard each of the 5 rumors neither believed in nor shared the claims. Using Fisher’s exact test, we conducted exploratory analyses to examine how belief and sharing behaviors were related. First, for the claim about the United States manufacturing the coronavirus to affect China’s economy, none who shared this rumor believed that it was true (*p*-value of 1 for Fisher’s exact test). In the case of the other 4 rumors, however, there was a significant association between belief and sharing (*p* < 0.001 for the rumors on drinking water and bat soup; *p* = 0.001 for the rumor on garlic, and *p* = 0.02 for the rumor on China creating the virus). Nonetheless, even with these 4 rumors, not all who propagated the rumors believed that they were true.

**Figure 2:**
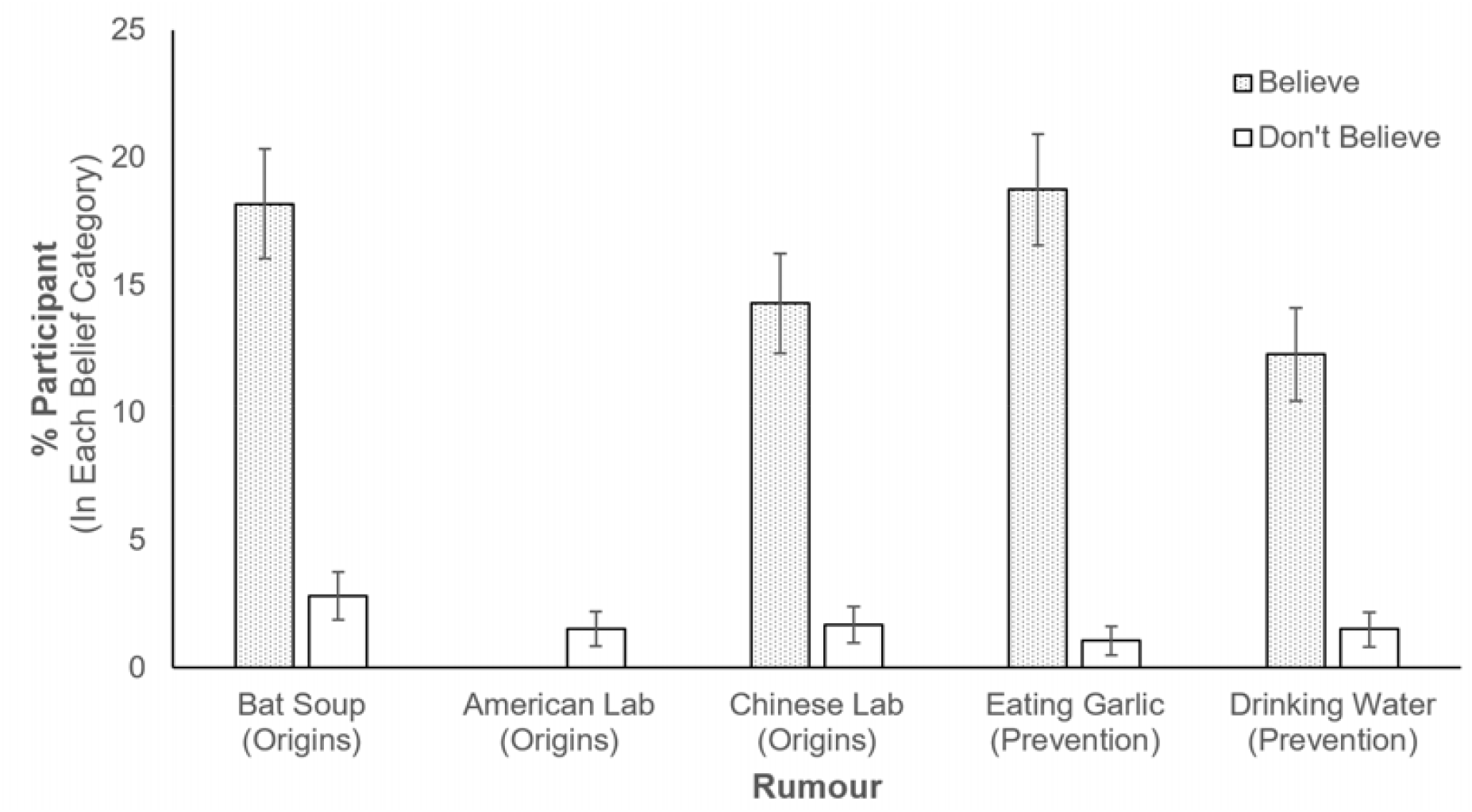
For each rumor, the vertical bar depicts the quantity of participants who shared each rumor, represented as a percentage of participants who believed or disbelieved each rumor. Vertical lines represent the 95% confidence interval.

Finally, Figure 3 depicts how participants had encountered COVID-19 rumors. As has been previously reported (Ippolito et al., 2020; Pew Research Center, 2018), social media platforms emerged as the leading sources, with 1 in 2 individuals reporting exposure through Facebook (55.5%) or WhatsApp (53.6%).

**Figure 3:**
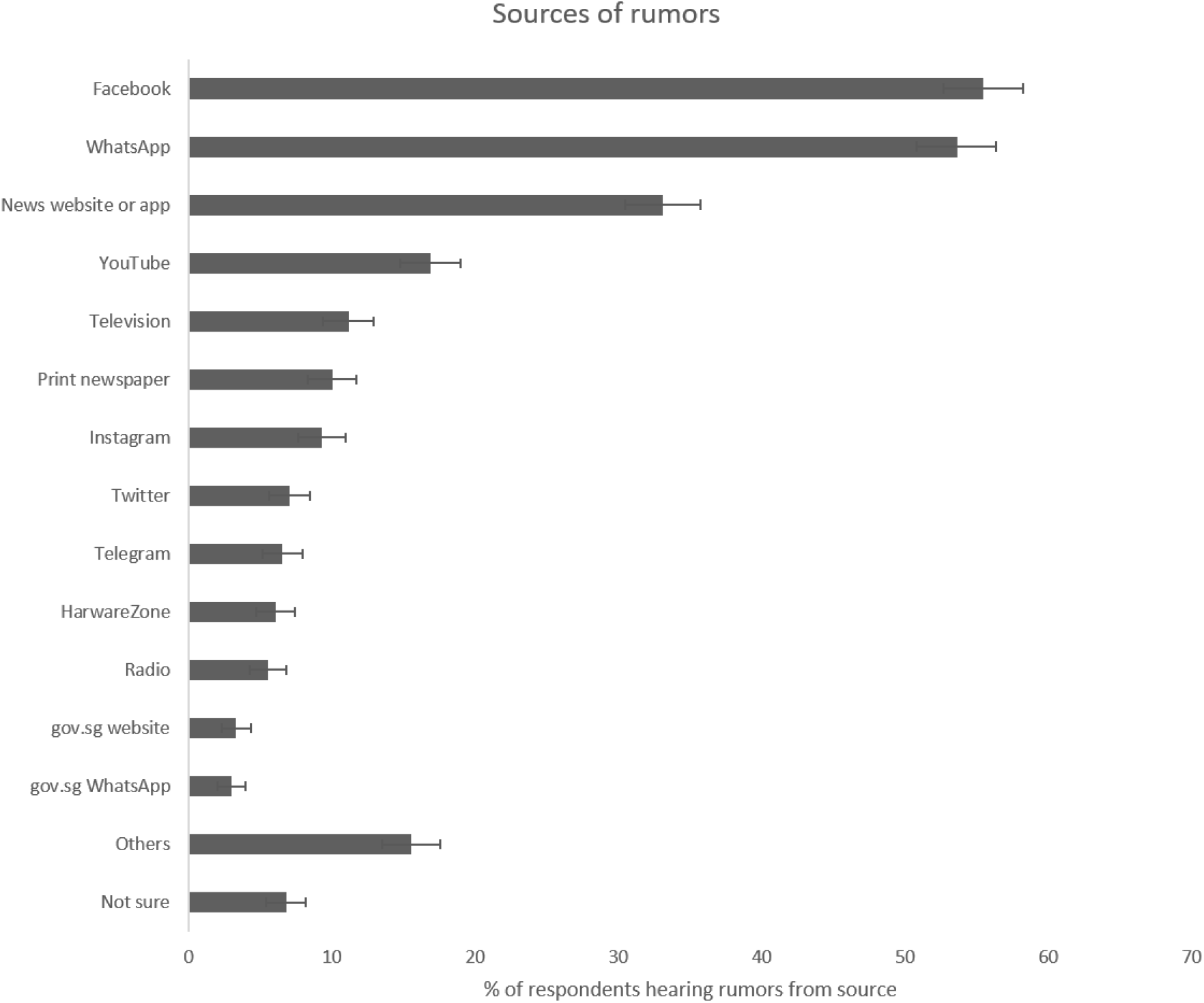
Sources where participants heard COVID-19 rumors from (with 95% confidence interval)

### 3.3 Predicting rumor hearing, sharing, and believing

Model 1 examined if any demographic or situational factors predicted the number of rumors heard. As shown in Table 2, participants reported hearing more rumors when confirmed local cases were few (early in the pandemic) (*b* = −0.621, *t*(1190) = −3.588, *p* < 0.001) or as lockdown restrictions were lifted (*b* = 1.129, *t*(1190) = 4.289, *p* < 0.001). Additionally, there was a trend for education to predict rumor exposure, with those with higher education hearing more rumors, (*b* = 0.077, *t*(1190) = 2.625, *p* = 0.009). (However, this association was not observed when Bonferroni correction was conducted.)

**Table 2.**
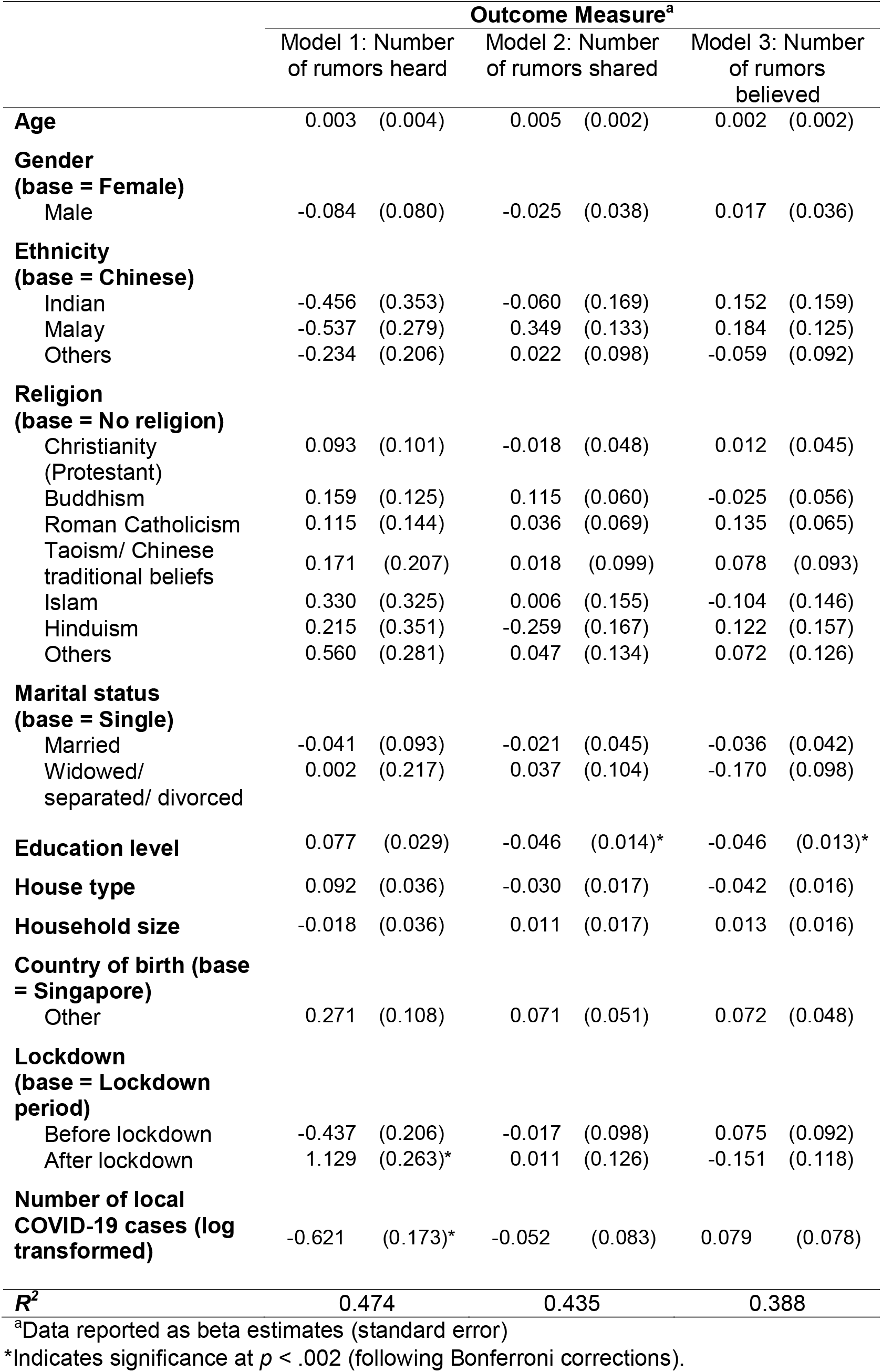
Predicting the number of rumors, heard, shared, and believed during the COVID-19 outbreak

|  | Outcome Measure<sup>a</sup> |  |  |  |  |  |
| --- | --- | --- | --- | --- | --- | --- |
|  | Model 1: Number of rumors heard |  | Model 2: Number of rumors shared |  | Model 3: Number of rumors believed |  |
| **Age** | 0.003 | (0.004) | 0.005 | (0.002) | 0.002 | (0.002) |
| **Gender (base = Female)** |  |  |  |  |  |  |
| Male | -0.084 | (0.080) | -0.025 | (0.038) | 0.017 | (0.036) |
| **Ethnicity (base = Chinese)** |  |  |  |  |  |  |
| Indian | -0.456 | (0.353) | -0.060 | (0.169) | 0.152 | (0.159) |
| Malay | -0.537 | (0.279) | 0.349 | (0.133) | 0.184 | (0.125) |
| Others | -0.234 | (0.206) | 0.022 | (0.098) | -0.059 | (0.092) |
| **Religion (base = No religion)** |  |  |  |  |  |  |
| Christianity (Protestant) | 0.093 | (0.101) | -0.018 | (0.048) | 0.012 | (0.045) |
| Buddhism | 0.159 | (0.125) | 0.115 | (0.060) | -0.025 | (0.056) |
| Roman Catholicism | 0.115 | (0.144) | 0.036 | (0.069) | 0.135 | (0.065) |
| Taoism/ Chinese traditional beliefs | 0.171 | (0.207) | 0.018 | (0.099) | 0.078 | (0.093) |
| Islam | 0.330 | (0.325) | 0.006 | (0.155) | -0.104 | (0.146) |
| Hinduism | 0.215 | (0.351) | -0.259 | (0.167) | 0.122 | (0.157) |
| Others | 0.560 | (0.281) | 0.047 | (0.134) | 0.072 | (0.126) |
| **Marital status (base = Single)** |  |  |  |  |  |  |
| Married | -0.041 | (0.093) | -0.021 | (0.045) | -0.036 | (0.042) |
| Widowed/ separated/ divorced | 0.002 | (0.217) | 0.037 | (0.104) | -0.170 | (0.098) |
| **Education level** | 0.077 | (0.029) | -0.046 | (0.014)* | -0.046 | (0.013)* |
| **House type** | 0.092 | (0.036) | -0.030 | (0.017) | -0.042 | (0.016) |
| **Household size** | -0.018 | (0.036) | 0.011 | (0.017) | 0.013 | (0.016) |
| **Country of birth (base = Singapore)** |  |  |  |  |  |  |
| Other | 0.271 | (0.108) | 0.071 | (0.051) | 0.072 | (0.048) |
| **Lockdown (base = Lockdown period)** |  |  |  |  |  |  |
| Before lockdown | -0.437 | (0.206) | -0.017 | (0.098) | 0.075 | (0.092) |
| After lockdown | 1.129 | (0.263)* | 0.011 | (0.126) | -0.151 | (0.118) |
| **Number of local COVID-19 cases (log transformed)** | -0.621 | (0.173)* | -0.052 | (0.083) | 0.079 | (0.078) |
| **R<sup>2</sup>** | 0.474 |  | 0.435 |  | 0.388 |  |

<sup>a</sup>Data reported as beta estimates (standard error)
\*Indicates significance at $p < .002$ (following Bonferroni corrections).

In Models 2 and 3, the same set of predictors were used to predict the number of rumors shared and believed, respectively. For both these models, those who were more educated shared or believed fewer rumors (Model 2: *b* = −0.046, *t*(1190) = −3.289, *p* = 0.001; Model 3: *b* = −0.046, *t*(1190) = −3.488, *p* = 0.001).

## 4. Discussion

During times of crisis, rumors have the potential to transmit misinformation and induce anxiety (Jin et al., 2014; Tran et al., 2020). Against the backdrop of the COVID-19 pandemic, we thus documented how individuals in the community came to receive, believe in, or share specific COVID-19 rumors.

First, we observed that rumor exposure was endemic. Nearly all participants had heard at least one rumor and were familiar with an average of 3 out of 5 popular claims assessed. Additionally, most rumor transmission occurred via social media channels (e.g., Facebook and WhatsApp), as others have noted (Islam et al., 2020; Wang et al., 2019).

Extending previous research, we further described how the base rate of believing or sharing rumors was far lower than the rate of exposure (with an average of <1 rumor believed or shared). Notably, belief and sharing behaviors did not always co-occur. In the extreme case of one particular rumor (that the COVID-19 crisis had been manufactured by the United States), not a single participant who reported forwarding the rumor actually believed in it. Although belief and sharing were linked for the other rumors we assessed, there continued to be – in each case – individuals who shared rumors without believing their veracity.

Our finding that COVID-19 rumors were disseminated even when disbelieved highlights the sheer difficulty of managing the so-called ‘infodemic’. Although similar findings had been reported outside the COVID-19 context (Chua and Banerjee, 2017), the World Health Organization and individual governments continue to issue – as the prevailing strategy – fact-checking statements to debunk rumors (Wong et al., 2020; World Health Organisation, 2020; Zarocostas, 2020). Our results bring to question the utility of such statements, since individuals continue to share claims despite perceiving them to be untrue.

Based on our findings, an alternative strategy may be to target individual vulnerabilities instead of rumor content. Given that rumor exposure changed with pandemic severity (e.g., the number of cases) and most individuals had encountered COVID-19 rumors, the ensuing question is why only certain individuals fell prey to either belief in or the sharing of these rumors. In our analyses, we found that educational level was a consistent predictor of vulnerability: although higher education predicted that an individual would hear more rumors, higher education was nonetheless protective, associated with fewer rumors shared or believed in. Consequently, it may be profitable to increase public awareness of knowledge and skillsets associated with higher education – for example, epistemology or scientific thinking (Chong et al., 2020; Chua and Banerjee, 2017). We note, however, that the correlational nature of our dataset precludes causal inferences, and further research will need to examine the efficacy of such strategies in curbing pandemic-related rumors.

### 4.1 Limitations

In describing these findings, we highlight several limitations of our research methods. First, we relied on participants’ self-reports regarding rumor exposure and behaviors. Although this strategy provided individual-level information (e.g., beliefs, demographics) not available in studies of actual rumor posts (e.g., when Twitter posts are mined), the survey method is vulnerable to recollection and reporting biases. Moving forward, future studies may opt to integrate digital documentation of rumor posts alongside self-reported measures.

As a second limitation, we only sampled rumors that were not time-sensitive. Given the limitations of the survey methodology, we could not track rumors that arose from fast-changing events on the ground – for example, rumors about the first COVID-19-related death in Singapore, or rumors about the availability of face masks (Asokan, 2020; Ministry of Communications and Information, 2020). It thus remains to be seen whether our findings can generalize to these forms of rumors.

### 4.2 Strengths

These study limitations need to be viewed alongside the putative strengths of our research methodology. To the best of our knowledge, our study represents the first attempt to identify individual vulnerabilities in the spread of COVID-19 rumors. The research involved a large sample size (1237 participants), captured pandemic-related dynamics over a long duration (5 months), and examined specific rumors that had been widely disseminated.

### 4.3 Conclusions

In conclusion, our study revealed that educational level was a protective factor amidst an onslaught of COVID-19 rumors. At a time when information regulation is crucial to resilience and well-being (Abdoli, 2020; Garfin et al., 2020), our findings provide a basis to manage the spread of rumors. In other words, it is not apparent veracity that makes a rumor ‘have it’. Instead, COVID-19 rumors are shared even when disbelieved, but may be stemmed through higher education.

## Data Availability

Data will be made available at request to the authors.

## Notes

### Competing Interest Statement

The authors have declared no competing interest.

### Clinical Trial

NCT04305574

### Funding Statement

This research was funded by a grant awarded to JCJL from the JY Pillay Global Asia Programme [grant number: IG20-SG002].

### Author Declarations

Yale-NUS College Ethics Review Committee (#2020-CERC-001)

